# Crisis Readiness and Innovation for Burnout Prevention Among Community Health Workers

**DOI:** 10.1101/2024.04.16.24305674

**Authors:** Ramya Raghavan

## Abstract

The performance of community health workers is an important factor in the success of health systems in underserved, vulnerable populations. Community health workers (CHWs) face work-related burnout. This study used a survey instrument to identify facilitators and barriers to a health crisis or pandemic preparedness. Causes of burnout and factors of crisis readiness were examined, including CHWs’ health status, readiness to use technology, and organisational culture. Findings suggest that more technology training and organisational support are needed to develop a resilient healthcare system. Increased workloads and demanding work schedules contribute to burnout, highlighting the need for support resources to promote the physical and mental health of CHWs. The study also collected data on CHWs’ perspectives on organisational commitment and individual innovation behaviours. The sample population is motivated to try new ideas and work methods to meet dynamic challenges and work expectations. Despite systemic shortcomings, more than 50% of them can envision a career with their community health organisation. Therefore, collaborative efforts are needed to create a work environment that promotes the physical well-being and professional development of CHWs. It is of practical importance for managers to value fostering innovation and organisational commitment in their healthcare workforce. In addition, our study is relevant to policymakers at local and central levels to plan national programmes for crisis preparedness and building a healthy nation.

## 1. Introduction

The COVID-19 pandemic has made it more important for community health workers (CHWs) to deliver key health interventions and promote health equity by improving outcomes for underserved populations and taking on important public health roles. In India’s rural health system, the main frontline workers are primary health centre (PHC) employees, auxiliary nurses (ANMs), accredited social health activists (ASHA), and Anganwadi workers (AWWs). The rural health sector of India is composed of policymakers in central and state government, doctors, nurses, pharmacists, health officers, ANMs, ASHA, PHC health workers, and AWWs. For ASHA’s outstanding contribution to protecting and promoting health, they were awarded the WHO director-general’s global health leaders award on 22 May 2022 [1]. PHCs are the means to meet national and social obligations of primary health care for rural communities, promote household economic well-being through improved health, reduce the risk of disease among breadwinners, reduce the cost burden of secondary and tertiary care, and increase confidence in the health care system [2]. While access to health services is an important focus, other administrative and human factors, as well as physical and digital infrastructure, play an important role in determining health and its delivery[3].

Prioritizing the mental health and well-being of health workers is an urgent global public health priority. Studies have shown that motivated human resources are key to improving performance and retaining health workers[4]. CHWs who are motivated, engaged, and satisfied with their work can improve HIV care and other interventions [5]. Health Workers work on the frontline, in rural regions with limited access to health resources and significant barriers to well-being. In villages, the health burden is disproportionately high, so it is important to know whether CHWs are prepared to address the challenges at work. Policies that recognise and support CHWs have the potential not only to improve rural health but also to reduce health inequalities [6]. Data shows that CHWs receive psychosocial support [7], recognition from communities and authorities, and interprofessional collaboration [8]. In addition, faith-based self-regulatory behaviours acted as resilience mechanisms for frontline health workers during the Covid-19 pandemic [9]. A better understanding of the work demands and motivational influences on health workers can help improve crisis preparedness and overcome health worker burnout. Community-based health research in India focuses on how ASHAs are trained and how they can be better equipped to help people in rural areas access health services. Most of these studies come to positive conclusions with few negative or mixed results, but they also reflect broader system constraints [10] [11]. This work is designed to develop an understanding of the rural health sector workers from primary health centres.

### 1.1 Problem Statement

There is an urgent need for the National Rural Health Mission of the Government of India to be prepared for another health crisis or pandemic. To improve health equity in rural areas, it is necessary to instill a sense of participation and organisational commitment among CHWs. Given the complex nature of this issue, rural PHC workers and volunteers represent an ideal sample population. What can help CHWs improve their work outcomes is a broad question of our interest. Studies have identified sources of motivation for CHWs at the individual, family, community, and organisational levels, but lack attention to psychological factors that influence CHWs’ motivation and job satisfaction[12]. This information is relevant to CHW burnout, and employee retention issues in the health sector [11]. Are our CHWs ready to innovate to address dynamic challenges at work? Research on organisational commitment and the use of innovative technologies among clinical health workers, demonstrate its effectives in the hospital setting [13]. There is a gap in this topic in the rural public health system. The answer sought will add knowledge to help make effective decisions about the increased financing and coordination to train, motivate and retain rural CHWs in low-income settings [14]. This study will explore individual perspectives on the work challenges faced as an up-to-date status on crisis preparedness. Our efforts in filling the research gap on crisis preparedness and individual innovation behaviour can benefit rural health organisations to build a healthy nation.

### 1.2 Hypothesis

Against this backdrop, this study used a survey instrument to identify facilitators and barriers to crisis readiness or pandemic preparedness. The present study aimed to address the following research question: Do rural CHWs experience work-related burnout? What are their views on crisis readiness variables and motivation for innovation?

The hypotheses developed are

- The challenges faced with digital technology readiness, and/or organisational work culture cause burnout among the sample population
- Fostering individual innovation behaviour, organisational commitment can help prevent burnout among the sample population

The study also tried to identify occupational health strategies needed in 2023 to prevent burnout. The data collected will benefit community health policymakers as they explore what can help CHWs improve their work outcomes in the context of crisis preparedness.

## 2. Literature Survey

Motivational influences on community health workers can help improve their work performance and overcome health worker burnout. Burnout is the most common mental health condition suffered by CHWs. Burnout is associated with emotional exhaustion, depersonalisation, and a diminished sense of fulfilment, which can affect their competence and health service delivery [15]. During the Covid 19 pandemic, much rural Community Health Workers (CHWs) put their safety on the line and travelled to villages to attend to patients. Health workers faced hostility, anger, and even death threats, as well as health misinformation, and dwindling trust in medicine and public health recommendations[16]. Key factors that could have triggered psychological distress among healthcare workers who were engaged in the management of COVID cases in India have been identified[17]. Poor investment in public health infrastructure, moral injury to provide terminal care for covid patients, long hours and inadequate support are some of the causes of psychological distress [18]. More research is needed to identify strategies to prevent burnout in addressing the future health crisis.

### 2.1 CHWs Nutrition and general health

Workplace health promotion involves policies, and environmental support to improve health, lifestyle, and behaviour [19]. CHWs with higher workloads and responsibilities are at a high risk of unhealthy lifestyles and mental health problems. CHWs should be given time and space at work to eat their homemade meals, drink water and follow healthy choices [20]. Making it easier for people to follow a healthy lifestyle by removing barriers at the personal, environmental, and system levels helps promote a healthy workforce [21]. CHWs to achieve adequate nutritional status, is crucial for coping with occupational stress. To fill the research gap on CHWs’ occupational experiences with a focus on physical well-being, self-report measures of body mass index and prescribed medications were collected. Masento et al. (2014) suggested that adequate water consumption may improve cognitive performance and mental well-being[22]. To highlight the important role of nutrition in mental health, this survey includes questionnaires on habitual lifestyle, including food consumption, eating patterns, water intake, and rest times, which may change due to workplace stressors, affecting immune response, and sleep [23].

### 2.2 Burnout causes

Community health workers under the Government of India’s National Health Mission are lauded as ’Covid Warriors’. However, it also draws attention to improving resources and tools such as budget structures and reports. to plan, manage and monitor utilization. A closer look at the impact of the Covid 19 pandemic on CHWs reveals that ASHA workers are struggling with anxiety and job insecurity and are constantly worried about their future [24]. This shows that management decisions can be improved through clear communication between ministries of health, ministries of finance, local government authorities, communities, health service providers, and CHWs. This provides the opportunity to make resource allocation decisions at the local level according to needs, disease patterns and priorities. In a systematic review, areas for collaboration were categorised as motivation, capacity building, relationship cultures, resource mobilisation and administrative support. and external factors [25]. This report shows that women involved in community care need help and that action should be taken to improve their situation. There isn’t enough research on what CHWs know, how they feel and what prevents or encourages them from working in the post-COVID-19 period. Do CHWs in rural areas suffer from burnout and stress that demotivates them to perform their tasks? The items in the questionnaire on burnout cause that affect work readiness were based on similar studies that explored what might worry CHWs today and in the future [26].

### 2.3 Crisis Readiness

There is literature on CHW performance, training and capacity building, but one-third of the articles were on specific projects focusing on the health situation. The review articles mention aspects of crisis preparedness such as timely preparedness, vulnerability or fear of infection, unsupportive management and high workload that can lead to stress and burnout. The plethora of pandemic-related reports identify several gaps in knowledge on how best to balance CHW organisational commitment and maintain health worker well-being[27]. Worksite health promotion approaches for better mental health, and nutrition has been recommended as an effective solution[21]. Recent evidence of the impact on health worker burnout in India suggests emerging new policies and guidelines may improve occupational resilience, and CHW occupational commitment [28] [29]. The global theme of remedial action to improve occupational resilience was represented by staff training, adequate equipment and supplies, and improved organisational support. Yet, there is limited evidence on the role of CHW programmes in promoting health workers’ technology skills and organisational commitment. We explored the current state of affairs through respondents’ perspectives on technological challenges, organisational work culture, and their impact.

#### 2.3.1 Organizational Work Culture

To support CHWs in carrying out their roles and responsibilities, it is important to use appropriate practise-based training methods and to match expected roles and responsibilities. There is an ongoing discussion on how public health systems can support health worker’s professional development [30] [31]. Country-specific literature reviews from Brazil[32] South Africa[33] and Botswana[34] echo similar themes. The staff development programmes in these reviews highlight gaps in policy to incentivize CHWs to provide health services to underserved communities. The retention issues project a global shortage of health workers. The reviews also show that CHW programmes need to be integrated into the community to be responsive to their needs. There is a need for integrated health services well-resourced, and well-monitored, to communicate the needs of village people to professionals and decision-makers. Another published study suggests that organisations must develop a work culture that focuses on their frontline workers [35]. To decipher the present status, the study questionnaire captured respondents’ views on incentives for CHWs, their working hours, career development training for hiring CHWs, and interactions with supervisors as indicators of organisational culture.

#### 2.3.2 Digital Technology Readiness

Stakeholders can make better decisions when communication channels from the government to local people and health workers are easy and accessible. Health workers who tried a telemedicine tool felt that it could improve their ability to interact with the patient [36]. The collaborative project in Ireland’s island population [37], aims to improve health through novel technological interventions in under-resourced settings. Such initiatives stop at the pilot stage due to a lack of training to use the digital application tool correctly and provide feedback about poor network coverage, and lack of infrastructure including access to electricity [38]. Opportunities for collaboration were grouped as motivation, skills, relationships, money, and support. Future work should explore the inclusion of mHealth technology as an important component in shaping population health [25].

Frequent training practises being needed for Indian CHWs to deal with phone applications. The apps like ASHApay, FP-LIMS, update data or make collections to manage health care financing and equitable distribution. We want to know if Indian CHWs in rural areas have the digital infrastructure, and knowledge and are ready to deliver mHealth in remote areas Emerging themes report challenges in infrastructure or technology support and usability of applications to manage daily tasks [37]. So we selected the scale for collaboration technology readiness which was originally developed in 2000 [39]. The questionnaire included indicator items about infrastructure or technology challenges in distance framework [40]. Overall, this study will add to existing knowledge, for crisis-related implementation, measures with digital technology and innovations to meet new expectations.

### 2.4 Individual Innovation Behaviour and Organizational Commitment

It’s imperative to empower CHWs with coping skills, knowledge, and training. However, it’s also critical that they continue to be part of the organization so that they can develop a long-term relationship with the people they serve. The development of interventions aimed at reducing attrition rates and psychological burnout depends on understanding the mental health needs and organizational commitment of ASHAs [3]. Organizational commitment and innovation among CHWs have not been discussed much in research. However, research has shown that burnout makes employees less innovative and motivated[41]. This can lead to a lower commitment to the organization overall. There is mixed empirical evidence about which incentives retain ASHAs in service. Additionally, interventions that provide work resources can help build an engaged workforce committed to the organization [42].

Female CHWs who are expected to deliver services with poor infrastructure may use simple innovations to minimize some of the challenges they face. Fostering innovation and commitment can be requirements for career development. In the healthcare industry, fostering engagement, and innovation is tied to commitment [43]. There is limited literature on how this is related to CHW retention, in rural organizations. This gap is addressed in this research by investigating the CHW’s career goals, innovation behaviour, and organizational commitment.

## 3. Methodology

### 3.1 Questionnaire Design

There is a greater need to understand the motivational factors that might influence the retention and performance of CHWs. As the survey was planned, several discussions with academic language experts and community health officers were held to verify the overall research design. The survey instrument was translated from English to Kannada and back-translated by volunteers to ensure uniform content and vocabulary. Community health officers at a nearby PHC completed a pre-test. The main survey instruments (burnout causes, crisis readiness, individual innovation, and organisational commitment) adapted from the current literature were used to measure the conceptual model of the study. Data were collected from primary health centre staff and rural community health workers in the Chikkabalapur district of Karnataka. Survey responses were collected through participant conversations, phone interviews, and Google online survey links from December 2022 to April 2023.

To assess their adherence to diet and activity recommendations, information on their general health status was investigated. The lifestyle and food habits in the workplace were measured using frequency questions from previous reports [21] [44]. The survey had five items from the technology readiness scale for distance framework. The ability of CHWs to complete daily tasks using digital technology through the frequency in which they can find plug points /electricity, time to understand how to do/ learn new apps/technologies, recent changes or difficulty with updates, slow internet speed /no signal were used to develop our questionnaire [40] [36]. Health agencies should focus on them to develop a positive organizational work culture as it has a direct effect on job satisfaction and staff attritions [45]. This study looked at respondents’ views on present organizational work culture (Figure 3.1) as it is a prerequisite for crisis readiness. We propose that community health workers who receive crisis readiness training and support will experience lower levels of burnout compared to those hospital employees who training[46].

**Figure 3.1:**
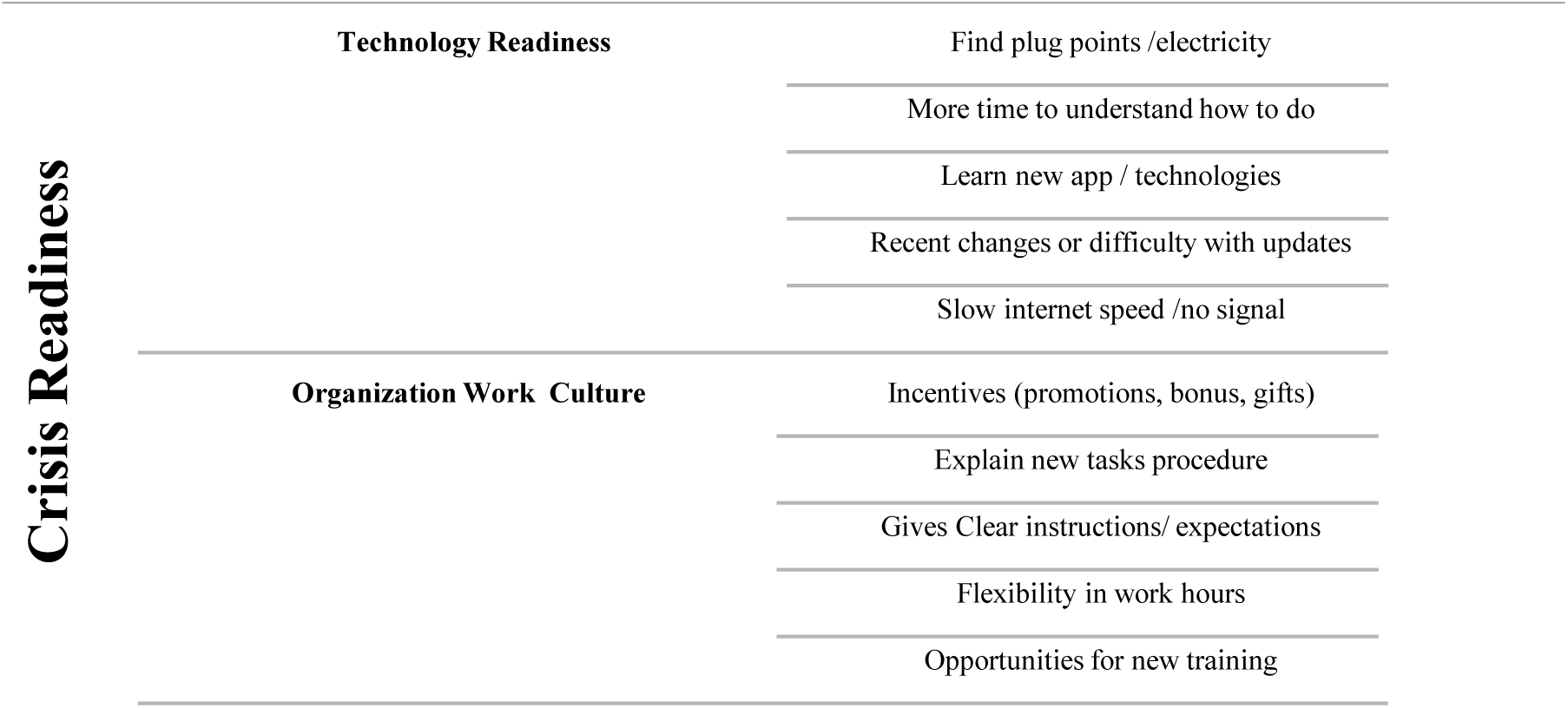
Crisis Readiness Factors and Questionnaire Statements

Burnout factors can affect the motivation and satisfaction of healthcare providers [47] at family, social, and community levels. We used the most commonly cited burnout causes (fear of infection inadequate supplies, increased workload, longer working hours, performing duties without proper training, and lack of information for new functions, procedures, and drugs)to work adapted from literature reported [26] [4]. Figure 3.2 shows the questionnaire items on [48] subscale, on the frequency of individual innovative behaviour [49] [50]. The scale had five statements (create new ideas to solve problems in my job, find new working methods or techniques, investigate ways for work efficiency, promote novel ideas so others might use them try out new ideas in my work). The organizational commitment, a scale developed [51] five items from the affective components of continuance commitment that were used in this study.

**Figure 3.2:**
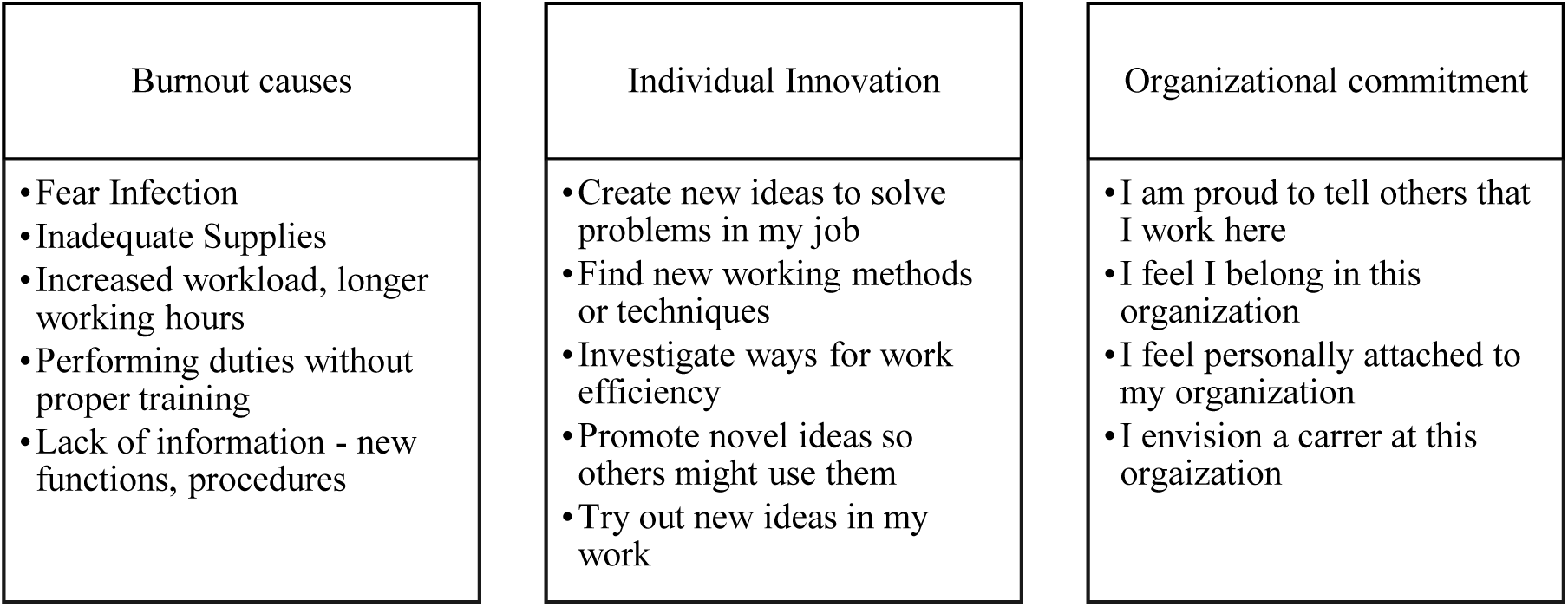
Questionnaire items for burnout, innovation and commitment

The study employed a survey method with the following schedule: A debriefing statement, demographic information requested, disclosures, and voluntary consent details. A set of survey questions were used to assess participants’ accounts of work-related experiences, and the details of their health and employment status. Each item is ranked on a 5-point Likert scale with a frequency range suitable to the question. The use of a 5-point ordinal Likert scale in the survey is important because it enables the capture of the overall frequency while also being ethical in asking participants to recall work-life events in a less detailed manner. This also ensures confidentiality because the scale ranges from “none” to “frequently,” which allows participants to provide a general idea of the frequency, they have experienced without having to recall specific details. Toward the end of the questionnaire, the participants were again asked about the voluntary consent disclosure to choose to submit their viewpoints on the potentially sensitive study topic.

### 3.2 Inclusion Criteria of the study population

Female healthcare personnel providing services to institutions in rural communities i.e., within the boundaries of a village, was selected under the inclusion category of our study. The study was carried out in Karnataka state, India from December 2022 to April 2023. We employed a mixed-methods approach of phone interviews, focus groups, and surveys (google Forms) through online WhatsApp and e-mail. The participants recruited were not given any compensation in money or kind. All study procedures were carried out by a team of women including the study’s principal investigator. To be included in our study, female participants had to meet the following inclusion criteria: (a) identify as healthcare workers from rural communities, (b) 18 years of age or older, and (c) presently pursuing employment at a primary healthcare centre. Sharing views or responses in the study is voluntary. The researchers distributed online links and paper survey forms to women who were involved in taking part in the study. A total of 50 healthcare workers completed our survey. The questionnaire was only given to adult participants voluntarily following an informed consent procedure.

### 3.3 Statistical Analysis

For each scale item in the survey instrument, descriptive statistics of Likert scale data were calculated (refer to appendix A3.3). Frequency tables and graphs were computed for data analysis. Exploratory factor analysis was performed and correlations in latent constructs were calculated. The reliability and validity values were checked for the latent constructs that had several questionnaire indicator items. Cronbach’s alpha, composite reliability and average variance extracted were used in measurement model reliability analysis. Discriminant validity was checked using the heterotrait monotrait ratios. All statistical analyses were conducted using Microsoft Excel, open-source JASP, (0.16.4 version) software, and R version 4.2.3 from the R Foundation for Statistical Computing Platform [52].

### 3.4 Demographics of the sample population

The survey collected responses from 50 participants aged 18–65 or more, 100% (50) were females. Out of these, 44% (22) of them are 35 – 44 years of age, 16% (8) have more than five, and 24% (12) have ten years of CHW job experience. 28% (14) were between 25 to 34 years old, and nine among them have been CHWs for five years or more. Table 1 shows there is a greater spread of educational levels among the sample population. 8% (4) of respondents attended high school, and 22% (11) did some skill-based occupational training after high school. 27 of 50 (54%) respondents have attended a degree college. The data on the educational level of the sample show that six of them have attended nursing college, and two are clinicians with advanced professional qualifications.

**Table 3.1:** Demographic information of the study sample population.

|  | Frequency | Percent |
| --- | --- | --- |
| **Age** |  |  |
| 18-24 | 11 | 22 |
| 25-34 | 14 | 28 |
| 35-44 | 22 | 44 |
| 45-54 | 2 | 4 |
| 65 and more | 1 | 2 |
| **Education Level** |  |  |
| MBBS MD DO / Clinical related | 2 | 4 |
| Attended High school | 4 | 8 |
| Nursing Degree / Nursing related | 6 | 12 |
| Paramedic Diploma / Occupational degree | 11 | 22 |
| Attended a college | 27 | 54 |
| **Job Description** |  |  |
| ANM / health worker | 3 | 6 |
| ASHA worker | 29 | 58 |
| Doctor / Physician | 2 | 4 |
| Allied Health Professional | 9 | 18 |
| Nurse/nursing related | 7 | 14 |
| **Healthcare experience** |  |  |
| Less than one year | 2 | 4 |
| 1-5 year | 17 | 34 |
| More than 5 years | 14 | 28 |
| More than 10 years | 17 | 34 |

## 4. Results and Discussion

In 2023, there is renewed push for the development of PHC workforce performance and avoiding worker shortages. Therefore, we enquired about the barriers to CHWs’ willingness to work so it can help identify burnout factors that affect job performance. The survey instrument aimed to collect self-reported CHW’s perspectives about physical health, technology readiness, organizational culture, individual innovation behaviour and organizational commitment.

### 4.1 Lifestyle and Healthy Behaviours

Workplace health promotion strategies aim to prevent illnesses, strengthen health, and improve well-being in the workplace.

Closer inspection of the table demonstrates, among the sample population about 20% of them suffer from blood pressure issues, asthma, menstruation, and skin problems (Table 4.2). The disease progression status shows 6 (12%) are highly symptomatic and 70% of them use medication to manage their condition. The study also found that within the sample population, 11 people are underweight, and 29 people have normal weight. Figure 4.3 also shows 8 (16%) are overweight. 2 (4%) are obese.

**Table 4.2:** Prevalence of common medical conditions among CHW women.

|  | Diabetes | Blood pressure | Asthma | Anaemia | Menstrual issues | Skin | Other |
| --- | --- | --- | --- | --- | --- | --- | --- |
| Early-stage medications not prescribed | 4 | 9 | 5 | 5 | 5 | 5 | 5 |
| Mild symptoms medications taken |  |  | 3 |  | 4 | 1 | 3 |
| Moderate symptoms medication taken | 1 | 2 | 2 | 3 | 2 | 4 | 4 |
| High Symptomatic medication taken | 1 |  |  | 1 | 3 |  | 1 |
| Total count | 6 | 11 | 10 | 9 | 14 | 10 | 13 |
| Percentage | 12 | 22 | 20 | 18 | 28 | 20 | 26 |

**Figure 4.3:**
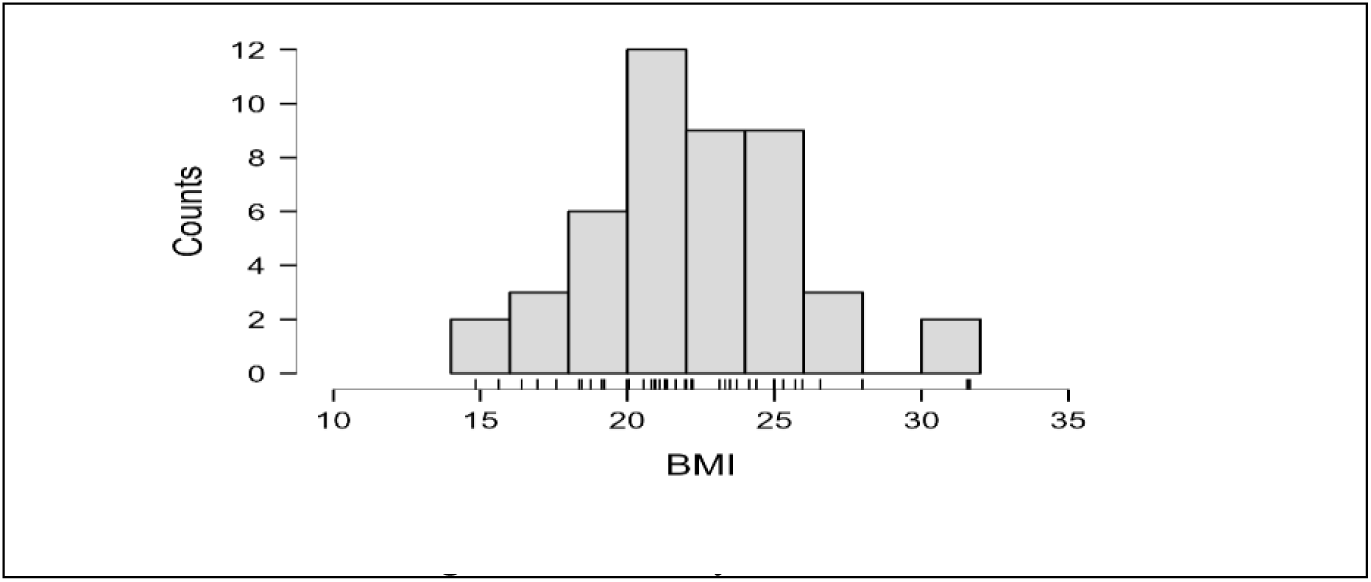
Body mass index

Calculating the frequency of lifestyle habits in the sample population is important because it can help identify trends and patterns. Figure 4.4 data suggests that CHWs skip meals, water intake, or rest breaks at work more often or frequently. Too much travel within on work shift can affect general health, as road infrastructure and transportation are poor in remote areas [53]. CHWs with higher workloads should be able to consume adequate food and water to avoid exhaustion. CHW’s adherence to healthy choices [20] reduces risk and mental health problems. The results exhibit the need for lifestyle resources for community health workers, nudging them to choose healthy behaviours [21].

**Figure 4. 4:**
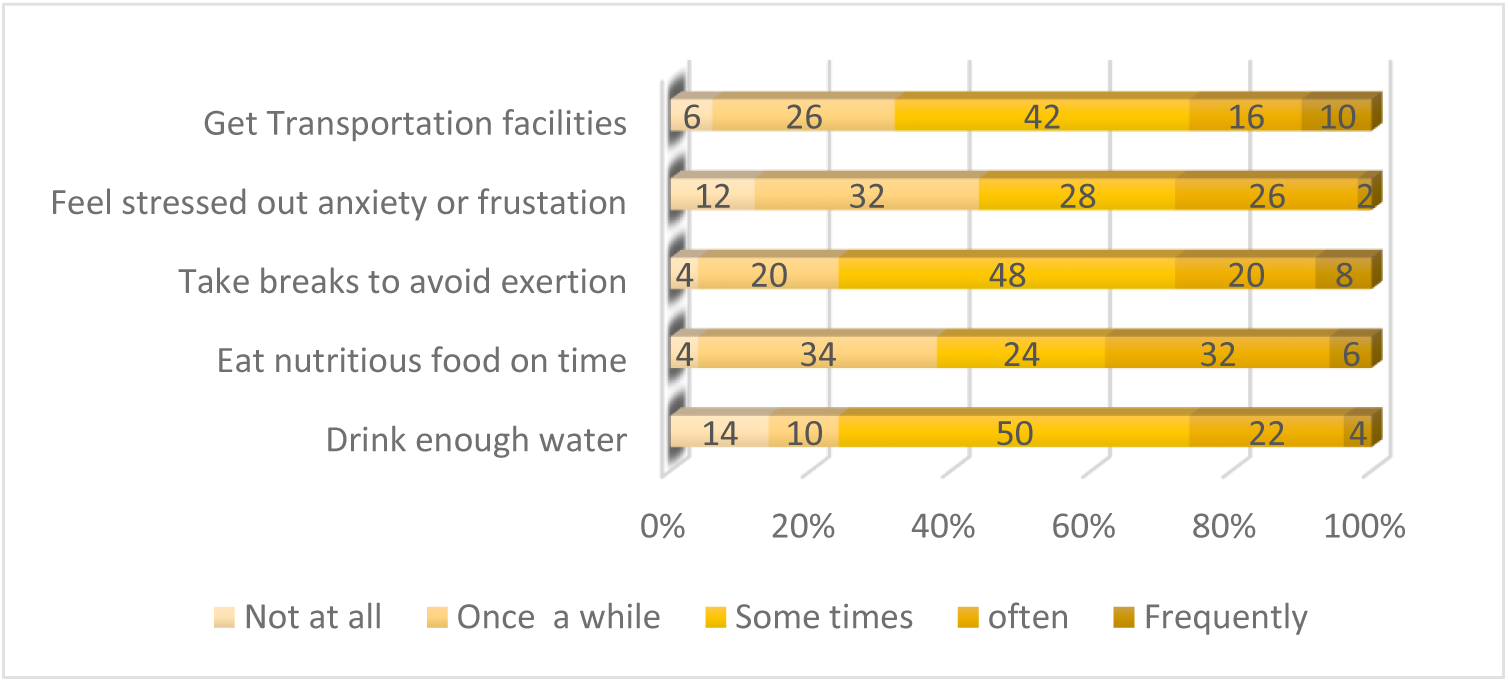
Frequency percentages on lifestyles-related questionnaire items

The results (Figure 4.4) show that CHW women in rural areas were indeed affected by anxiety, frustration at work and lack of proper transportation facilities for field work adds to their woes. It is evident from this table that pre-existing conditions, and lifestyle factors when added to stress factors at work, can increase the health risks [23].

Promoting mental health and well-being is essential, as anxiety and frustration-related burnout can contribute to a higher risk of health problems [3]. One argument for why health has declined is that of organizational work culture. Therefore it is important to address the personal and professional concerns of healthcare workers and provide them with training or coping skills for a positive occupational experience [7].

### 4.2 Burnout Causes that Affect Work Readiness

CHWs play a critical role in providing healthcare services to underserved communities, and barriers that impact their work can have significant consequences for the health outcomes of these communities [15]. It is important to check if healthcare workers in our sample population experience work fatigue or burnout and its impacts.

In this section of the survey, the respondent gave their level of agreement to statements describing burnout causes that affect their job outcomes adapted from Barqawi et.al 2021[26] in the form of the 5-item Likert scale. There is an increased strain on the CHWs when they face increased work and long working hours without proper supplies and training. There is a clear trend in Figure 4.5, 26% of CHWs face increased workloads and long working hours frequently. This was one of the factors that triggered psychological distress all around the world during covid infection. The present status reflecting a similar situation among healthcare workers in our respondents [17] implies worker shortages. Other factors that impact

**Figure 4.5:**
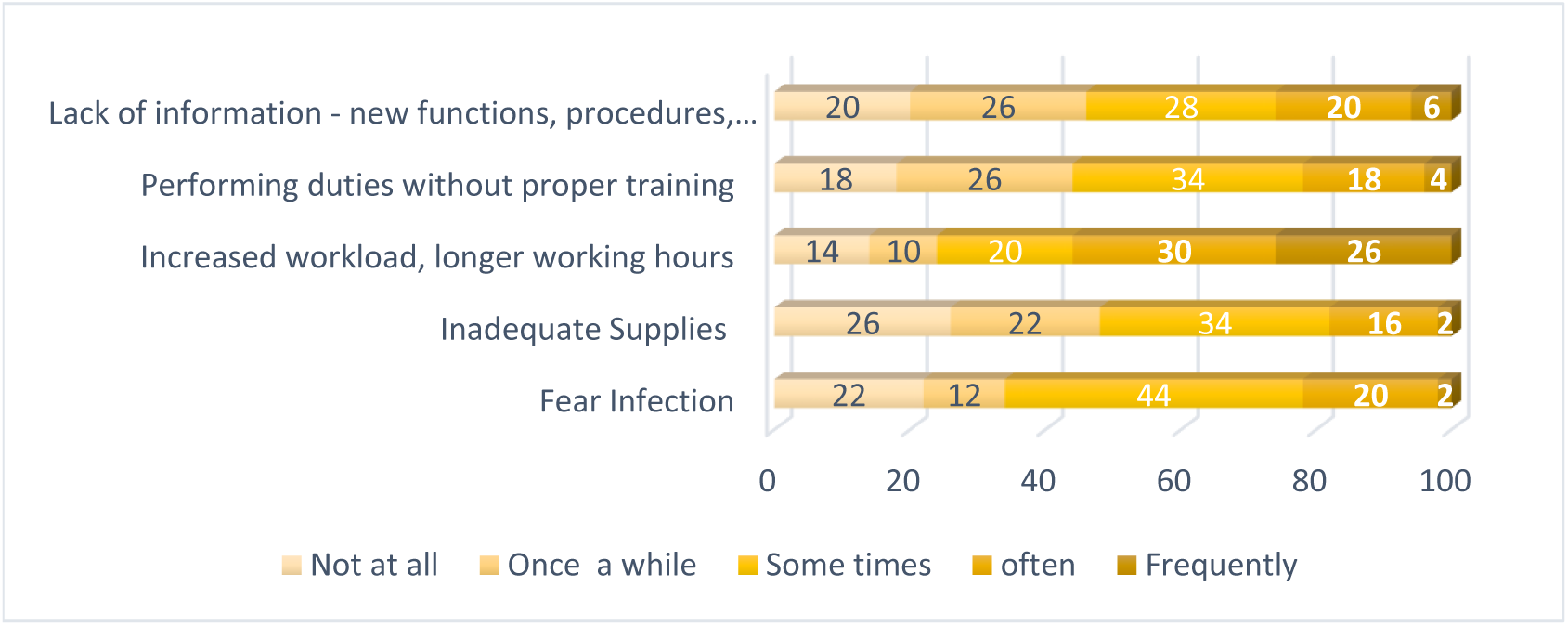
Burnout causes affecting willingness to work

CHW’s long working hours include a lack of training and support, inadequate resources, and transportation issues. Our data matches with similar reports on the heavy workload and long working hours[54].

Several recent reports show burnout-related issues can lead to poor quality of care, low job satisfaction, high turnover rates psychological distress [18]. Developing national, region-specific crisis readiness policies and updating, performance indicators is crucial to face other pandemic-like situations.

### 4.3 Crisis Readiness

One of the aims of this work was to find out if community health workers are provided with crisis readiness resources and support. Data were collected to find the present status of supporting organizational and infrastructure elements. To evaluate crisis readiness digital resources’ ease of use and questionnaire items in health workforce development and training was used.

#### 4.3.1 Organizational work culture and Training

CHWs impact community wellness and If CHWs are supported to realize their maximum capacity it will have a positive impact. About half of the CHWs (50%) report that they receive monetary incentives, such as compensation and benefits.

20-40% of the sample population report getting non-monetary factors, such as training, support, and CHWs no more hold on to the psychological burden of the pandemic [24]. One argument for why psychological issues have declined is shown in our results, that financial and non-financial incentives are used to improve the performance of community health workers There is now wide consensus that supporting organizational and infrastructure elements are lacking to succeed in public health work. Further, it suggests that there is a need to implement training and supervision, more often to achieve crisis readiness in the future [26]. In general, it appears that providing community health workers with crisis readiness training can help them feel more prepared and confident in dealing with unexpected situations, reducing stress and burnout [30] [31].

Different questionnaire items have been employed to support the hypothesis that there is a relationship between organisational work culture and the causes of burnout among community health workers. The results of this work add to the existing theme that CHW programmes are complex and a solid support system is needed to strengthen the CHW workforce [35]. The most likely reason crisis readiness is possible is when CHWs’ are considered stakeholders during the expansion and assessment of CHW-led projects [55].

#### 4.3.2 Technology Readiness vs Ease of Use

Digital technology readiness and collaboration are essential for effective primary health care. This means the stakeholders need to achieve a common ground and be prepared for intersectoral collaborative projects using digital technology. The survey enquired about technology readiness (infrastructures like internet, electricity, phone application tools, etc.), where primary health care data collected needs to be shared with several local health, state, and central government sectors. This study established that finding electricity sources and stable internet connections was challenging more frequently by 24% of the CHWs in fieldwork. Results show CHW technical competency tends to decline after training, as 34% of the population has difficulty operating mobile applications. It seems 22% CHWs require follow-up to learn new online forms or apps and 36% need multiple supervised practice sessions to keep pace with the newer app updates. Data suggest CHW performance may be improved by supportive supervision that focuses on quality assurance and problem-solving. Study participants agree with the existing literature and welcome the move toward digitization [36]. Despite the implementation challenges including lack of access to electricity [38], it has the potential to reduce the time and workload. However, several challenges faced are often invisible to policymakers. This observation is reported from low-income group countries, where there are limited resources to assist or use electronic documentation tasks [37].

Digital technology infrastructure allows everyone in the planning and delivery of health care to work together. More recent attention has centred on the provision that organizations can collaborate using digital information systems [56]. In India, resource allocation, and decision-making within the public health sector are made by the local government [57].

Results support engagement with all stakeholders for better acceptability and credibility of the CHW program [58]. In an in-depth Lancet review [59] authors recommend that the main source of funding for primary health care should be public resources, and it should be distributed fairly across different levels of service.

### 4.4 Individual Innovation and Organizational Commitment

With the current technological advances and increased performance expectations CHWs are facing more challenges and there is a need to find ways to overcome mental burnout more than ever before. As a possible remedy option, it is important to know if community health workers are willing to make innovations to combat challenges at work. The survey responses on innovation and commitment traits gathered support for existing knowledge that innovation behaviour can help overcome burnout by providing a sense of purpose and motivation [42]. The data shows 60% CHWs in our sample do create new ideas, and try out new ideas to solve problems and it could involve finding a new working methodology. As Figure 9 shows, more than 50% of individuals are motivated to find ways to work efficiently. Data shows, increasing innovation at work can help improve performance as well as efficiency[55]. Therefore, creative and innovative are critical solutions for employees against burnout-related challenges. The innovation behaviour data led us to the premise that CHWs are motivated to bring in innovation as they anticipate a long-term career and promising organizational work culture [60]. Another important aspect we investigated was the organizational commitment by using various indicator items. Figure 4.9, that the affective component of organizational commitment is desired among the respondents [43] similar to those of medical professionals in clinical settings. The respondents are very much motivated towards their work, this bolsters reports on lack of motivation, capacity building, and administrative support [25]. In contrast to earlier observations, due to their community engagement, CHWs remain positive in their work. Factors such as their strong sense of purpose, recognition for their efforts, support from their community as well as training and empowerment might sustain a motivated workforce [3] [7] [8].

**Figure 4.6:**
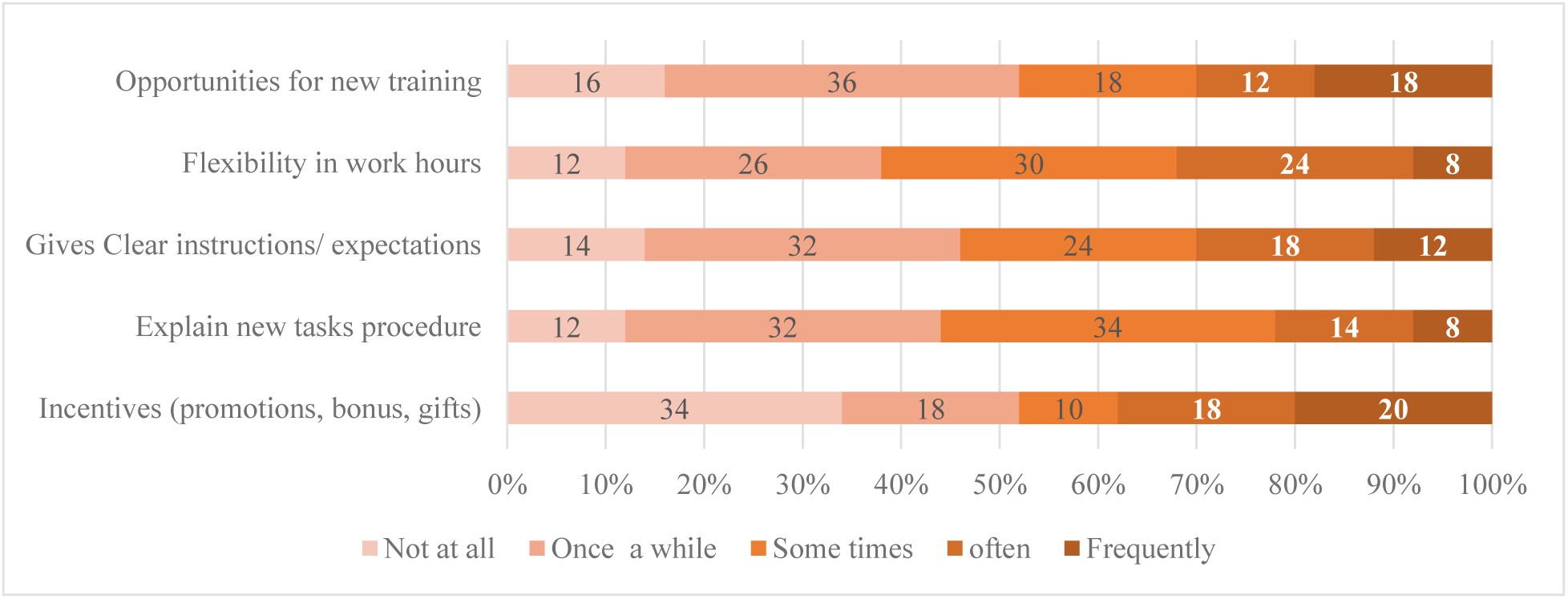
Organizational Work Culture Indicators

**Figure 4.7:**
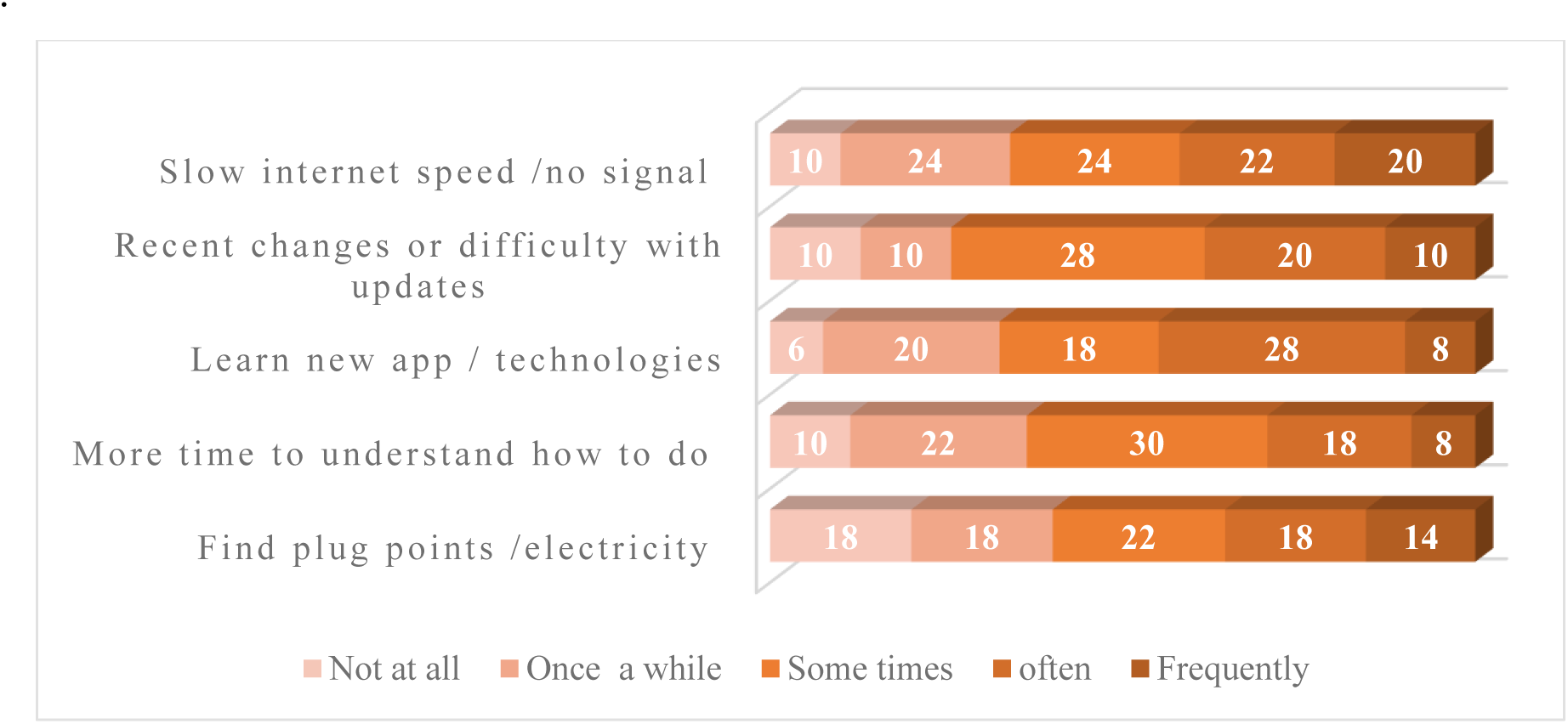
Technological challenges faced by CHWs Adapted from Caldeira et al. 2022 [40]

**Figure 4.8:**
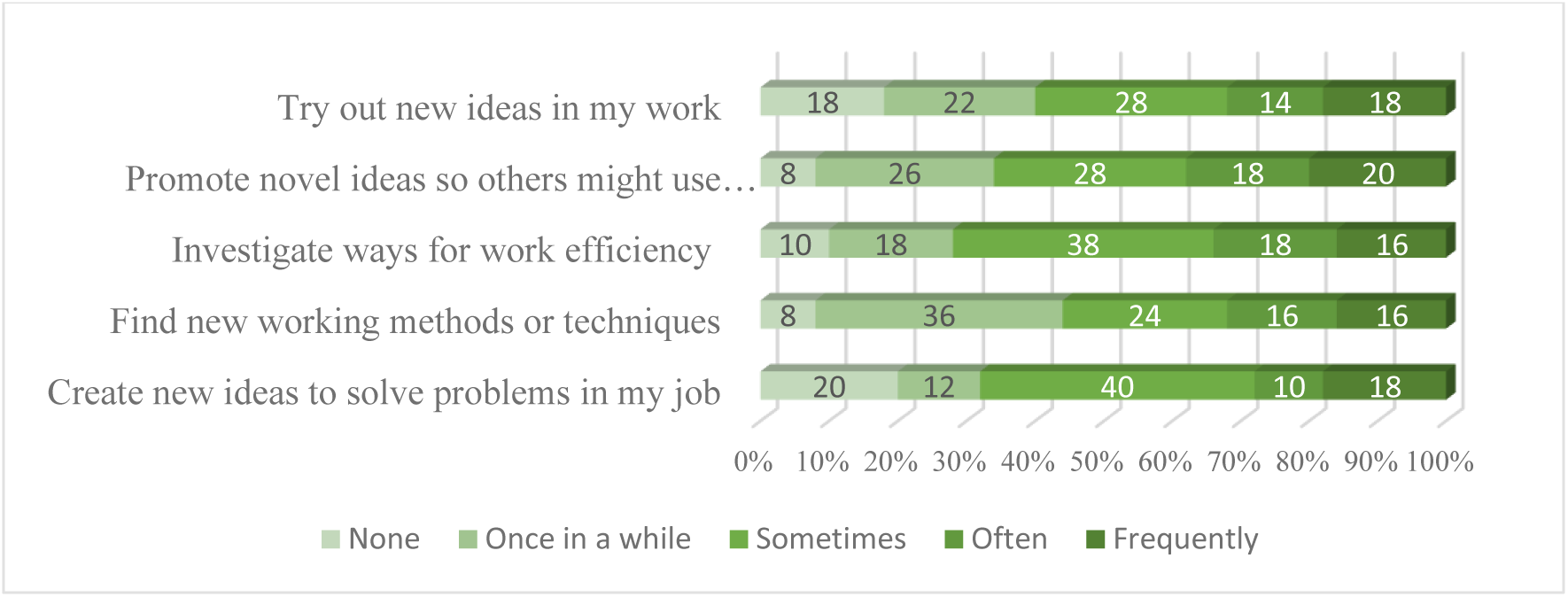
Individual Innovative Motivations

**Figure 4.9:**
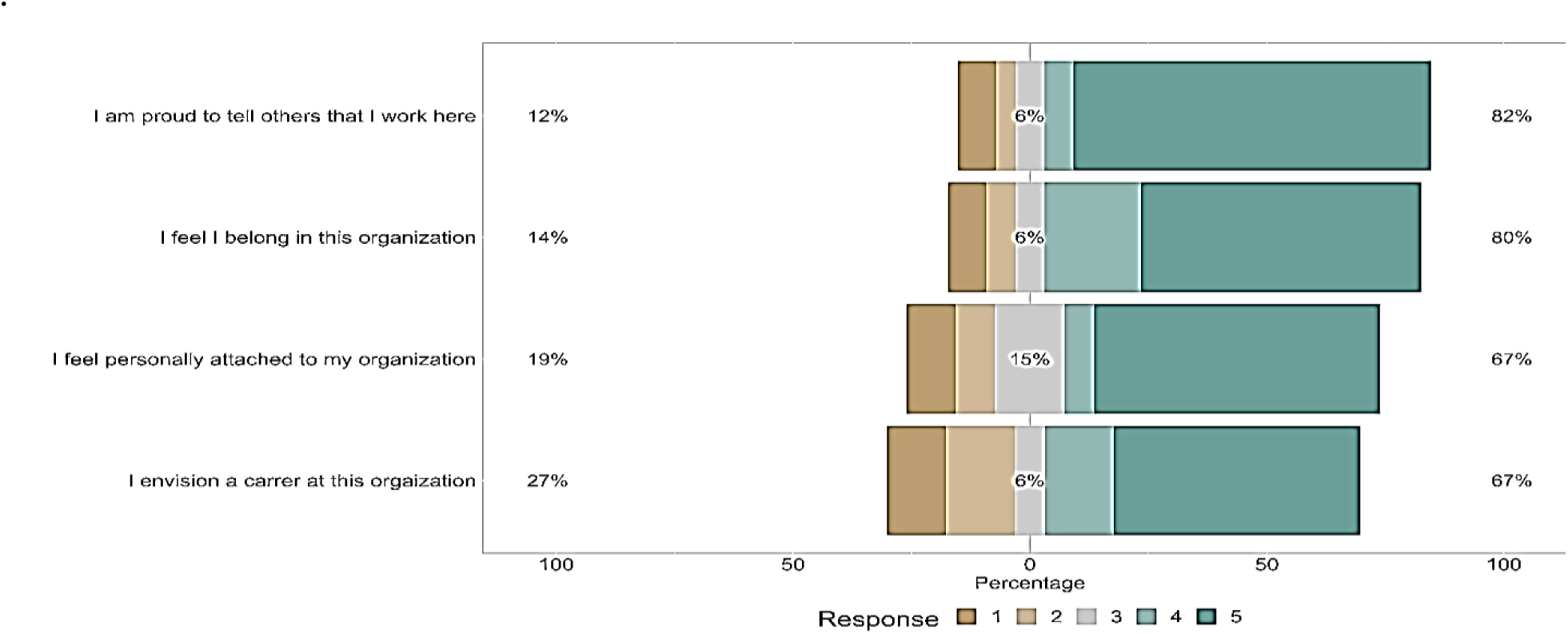
Organization Commitment Responses Likert scale: 1=Never 2= Monthly 3=Weekly 4=2-3 times a week 5 =Daily.

It provides insight into CHW’s perceptions of creating positive work commitment with the PHC [6]. Using the binomial test, we found 75.5% (p < .001) are proud to tell others about their employment. Similarly, 60% of the respondents feel they belong or are attached to the organization (p < 0.001). More than 50% of the CHWs envision a career with this current organization. This study has demonstrated support for the hypothesis that there is a direct relationship between CHWs’ affective organizational commitment, innovative behaviour, and burnout prevention. This challenges earlier reports on the hospital industry [10] [11] but adds valuable information to the existing literature on public health to foster innovation and the mental health of employees.

### 4.5 Exploratory Factor Analysis

Exploratory factor analysis (EFA), is conducted, to confirm a theoretical framework and identify data patterns and relationships. The factor analytic technique helps explain patterns of covariation that highlight the relationships by grouping the questionnaire. The indicator items are those observed or manifest (i.e., directly measured) variables using the Likert scale. The unobserved or latent constructs can be identified, as EFA allows to group of a large number of variables (observed variables) into smaller representative factors (latent factors). To help determine if the exploratory factor analysis is appropriate, Kaiser’s measure (KMO) of sampling adequacy, and Bartlett’s test correlation matrix has significant correlations were computed. A KMO sampling adequacy value above 0.5 (proportion of variance among variables) shows our data is suited for factor analysis. Bartlett’s Test of Sphericity (< 0.05) with an acceptable significance level (Appendix) suggests factor analysis is appropriate [61]. Using EFA analysis, five latent factors were identified, by promax oblique rotation that allowed the extract factors to correlate using the extract factor structure. The extracted latent variables supported our study framework thus validating the survey instrument and results obtained.

The model fit measures Root Mean Squared Error of Approximation (RMSEA) = 0.036, and Standardized Root Mean Square Residual (SRMR) = 0.04, which were less than 0.08. The Comparative Fit Index (CFI) and Tucker-Lewis Index (TLI) indices, result values were close to or greater than 0.95 suggest to reflect an acceptable model fit [62], and factors obtained from EFA were validated. The Chi-square goodness of fit statistics was not statistically significant, perhaps due to the small sample size.

EFA method finds factors that explain the correlations between variables. Five independent factors were obtained starting from many questionnaire items. Table 4.3 indicates the correlations between the factors. The factor correlations between burnout causes and technology readiness (0.419), and organizational work culture (0.184) suggest that inadequate technology resources do contributes to burnout among community health workers. Data also suggest a relationship between individual innovation behaviour (0.391) and burnout, (but not organizational commitment (0.074)). Due to study limitations and a small sample size, we cannot speculate much from the factor analysis. Nevertheless, it supported the hypothesis proposed. Structural equation modelling methodology does not require normally distributed data and bootstrapping allows us to get solutions with comparatively smaller sample sizes [52]. To identify the future course of action, we tested the variables: technology readiness, burnout causes, and individual innovation by partial least square modelling (Appendix table A4.5). This analysis using bootstrapping suggested that technology readiness, burnout causes (barriers) and individual innovation do relate to each other confirming our EFA analysis. In the future, more extensive studies with a higher sample size will be required to compute the relationships and their statistical significance. Table 4.4 presents the reliability indices for latent constructs that were checked as per the criterion given in the literature.

**Table 4.3:** Factor Correlations - Exploratory Factor Analysis.

|  | Burnout<br>causes | Orgn work<br>culture | Technology<br>readiness | Individual<br>Innovation |
| --- | --- | --- | --- | --- |
| Burnout causes | 1 |  |  |  |
| Orgn work culture | 0.184 | 1 |  |  |
| Technology readiness | 0.419 | 0.433 | 1 |  |
| Individual Innovation | 0.391 | 0.023 | 0.221 | 1 |
| Organization commitment | 0.074 | 0.279 | 0.21 | 0.247 |
| Additional fit indices |  |  |  |  |
| RMSEA | RMSEA<br>90% CI | SRMR | TLI | CFI |
| 0.036 | 0 - 0.083 | 0.04 | 0.956 | 0.978 |

### 4.6 Evaluation of Reliability and Validity Criterion

Frequentist scale reliability statistics for internal consistency of questionnaire items using Cronbach’s alpha (α) = 0.86 and, McDonald’s omega (ω) = 0.82, show good reliability (above 0.7) fit. In factor analysis, we used several latent constructs (figure 3.1, 3.2), with reflective measurement indicator items. This requires evaluating the reliability at the construct level. Summarized in Table 4.4, are reliability and validity values.

**Table 4.4:**
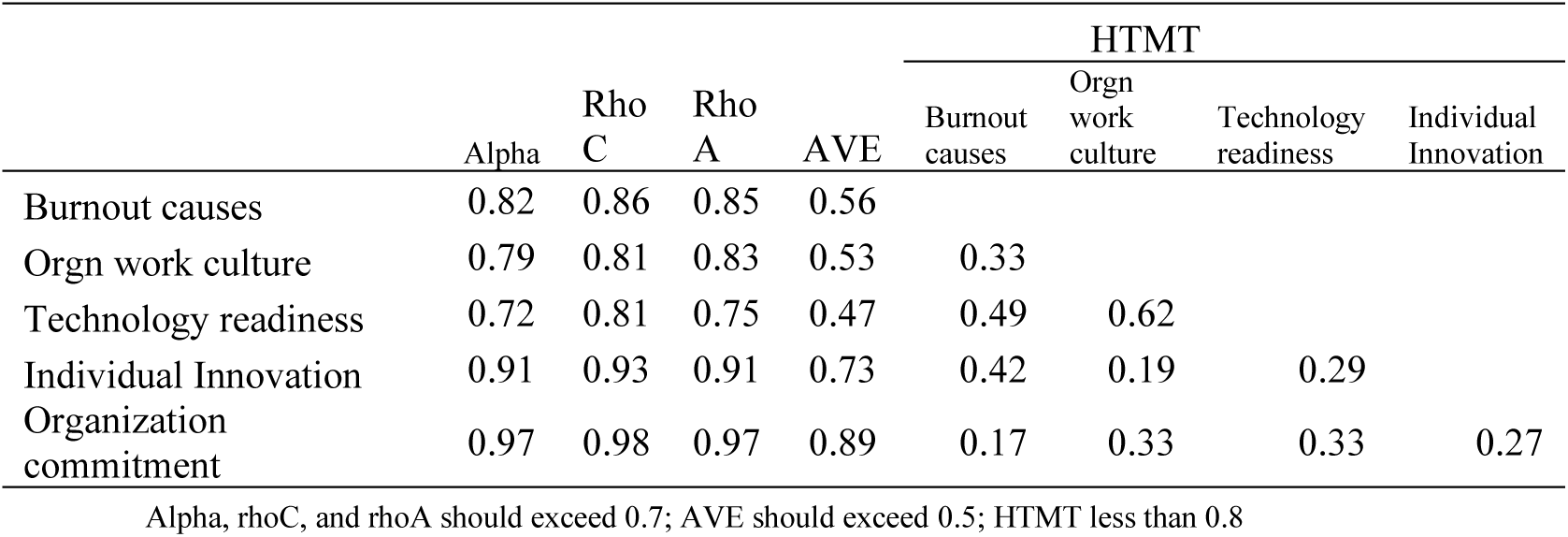
Evaluation of the Reliability and validity standards.

|  | Alpha | Rho<br>C | Rho<br>A | AVE | HTMT |  |  |  |
| --- | --- | --- | --- | --- | --- | --- | --- | --- |
|  |  |  |  |  | Burnout<br>causes | Orgn<br>work<br>culture | Technology<br>readiness | Individual<br>Innovation |
| Burnout causes | 0.82 | 0.86 | 0.85 | 0.56 |  |  |  |  |
| Orgn work culture | 0.79 | 0.81 | 0.83 | 0.53 | 0.33 |  |  |  |
| Technology readiness | 0.72 | 0.81 | 0.75 | 0.47 | 0.49 | 0.62 |  |  |
| Individual Innovation | 0.91 | 0.93 | 0.91 | 0.73 | 0.42 | 0.19 | 0.29 |  |
| Organization<br>commitment | 0.97 | 0.98 | 0.97 | 0.89 | 0.17 | 0.33 | 0.33 | 0.27 |

Alpha, rhoC, and rhoA should exceed 0.7; AVE should exceed 0.5; HTMT less than 0.8

The reliability of a survey scale refers to the degree to which the questionnaire items in the scale are measuring underlying latent constructs. The Cronbach alpha and Rho_A reliability criterion are suggested to be above 0.7. The composite reliability Rho C is above 0.80, yet below 0.95, which indicates appropriate composite reliability[62]. The reliability coefficient Rho A, Cronbach’s alpha, and the Rho C composite reliability are within the criterion range and acceptable [63].

The average variances extracted (AVE) in our results exceed 0.50 indicating appropriate convergent validity of the constructs. Almost all fit this except the technology readiness construct measured at 0.48 for average variance extracted which is moderate and acceptable. The heterotrait - monotrait ratio of correlations (HTMT) had been recommended to assess the discriminant validity of latent variables [64]. We found in our study the HTMT criteria are met, as values well below 0.90 would suggest an absence of any problems with discriminant validity [65]. The average variance extracted (AVE) for each construct, was a minimum of 0.50, thus demonstrating adequate convergent and discriminant validity [66] [67]. The evaluation of collinearity along the factors (less than 3.3), does not point to collinearity issues, but variance inflation factors VIF values do not apply to reflective measurement model indicators [63].

## 5. Conclusions

This study used a survey instrument to identify present challenges to pandemic preparedness. It supports our hypothesis that the challenges faced with digital technology readiness are a major cause of burnout among the sample population. The positive responses on motivation for innovation and organization commitment validate the hypothesis that fostering these qualities among the workforce can help prevent burnout. The difficulties reported with technology handicaps, longer work hours, and higher workload suggest improving the organisational work culture to deliver more frequent training and follow-up supervision. Non-monetary factors, such as training, support, and recognition, can also affect CHW motivation, performance, and job satisfaction, and should be considered in interventions to improve CHW retention rates. Our efforts to quantify how often CHWs are motivated to innovate to prevent workplace burnout have several theoretical and practical implications. It is of practical importance that managers place value on encouraging innovation and organisational commitment among their healthcare workers. It will be useful for policymakers at local and central levels to plan workforce development programmes or undertake efforts for crisis preparedness in India’s public health system.

### Limitations

This study is limited by its small sample size, this could imply that themes from these studies with smaller samples are not exhaustive. While it was preferable to conduct telephonic interviews, anonymity is perceived better in paper or online survey methods and avoids social desirability bias. The conclusions may have been oversimplified since the research variables did not examine all the crisis preparation supports offered to rural CHWs to help them deliver healthcare in rural regions. Additionally, this study data is limited to opinions of females, associated with primary health centers from Karnataka, India. Our work cannot be generalized to healthcare populations in other sectors.

## Data Availability

All data analyzed during this study are included in this manuscript

## Statements and Declarations

### Ethical Approval

This study was performed in line with the principles of the Declaration of Helsinki. The studies involving human participants were reviewed and approved by Sri Sathya Sai University for Human Excellence, Kalaburagi, Karnataka, 585313, India. The present study obtained consent from the Sri Sathya Sai University for Human Excellence, Kalaburagi, Karnataka, 585313, India ethics committee (Approved). The study protocol was approved and fulfilled ethical guidelines for human research.

### Compliance with Ethical Standards

Informed consent was obtained from every participant to be included in the study. The authors affirm that human research participants provided informed consent for the publication of their survey responses.

### Consent for publication

The authors affirm that human research participants provided informed consent for the publication of survey response data. This statement confirms that consent to publish has been received from all participants.

### Availability of supporting data

All data analyzed during this study are included in this manuscript (and its supplementary information files). The dataset generated will be shared on reasonable request to the corresponding author.

## Acknowledgments

The authors acknowledge the healthcare workers who contributed their time and experiences as respondents in this study. We would like to extend our gratitude to all the women who participated in this study, the research staff and volunteers, and community stakeholders.

Disclaimer: The content of this publication is solely the responsibility of the authors and does not necessarily represent the official views of any other authority or parties.

## Authors’ contributions

All authors whose names appear on the submission made substantial contributions to the conception or design of the work; or the acquisition, analysis, or interpretation of data;

